# The aerobiology of SARS-CoV-2 in UK hospitals and the impact of aerosol generating procedures

**DOI:** 10.1101/2022.09.07.22279662

**Authors:** Susan Gould, Rachel L Byrne, Thomas Edwards, Ghaith Aljayyoussi, Dominic Wooding, Kate Buist, Konstantina Kontogianni, Allan Bennett, Barry Atkinson, Ginny Moore, Jake Dunning, Stacy Todd, Marie-Claire Hoyle, Lance Turtle, Tom Solomon, Richard Fitzgerald, Mike Beadsworth, Paul Garner, Emily R Adams, Tom Fletcher

## Abstract

**Background:** SARS-CoV-2 nosocomial transmission to patients and healthcare workers (HCWs) has occurred throughout the COVID-19 pandemic. Aerosol generating procedures (AGPs) seemed particularly risky, and policies have restricted their use in all settings. We examined the prevalence of aerosolized SARS-CoV-2 in the rooms of COVID-19 patients requiring AGP or supplemental oxygen compared to those on room air.

**Methods:** Samples were collected prospectively near to adults hospitalised with COVID-19 at two tertiary care hospitals in the UK from November 2020 – October 2021. The Sartorius MD8 AirPort air sampler was used to collect air samples at a minimum distance of 1.5 meters from patients. RT-qPCR was used following overnight incubation of membranes in culture media and extraction.

**Results:** We collected 219 samples from patients’ rooms: individuals on room air (n=67), receiving oxygen (n=65) or AGP (n=67). Of these, 54 (24.6%) samples were positive for SARS-CoV-2 viral RNA. The highest prevalence was identified in the air around patients receiving oxygen (32.3%, n=21, CI95% 22.2 to 44.3%) with AGP and room air recording prevalence of (20.7%, n=18, CI95% 14.1 – 33.7%) and (22.3%, n=15, CI95% 13.5 – 30.4%) respectively. We did not detect a significant difference in the observed frequency of viral RNA between interventions.

**Interpretation:** SARS-CoV-2 viral RNA was detected in the air of hospital rooms of COVID-19 patients, and AGPs did not appear to impact the likelihood of viral RNA. Enhanced respiratory protection and appropriate infection prevention and control measures are required to be fully and carefully implemented for all COVID-19 patients to reduce risk of aerosol transmission.

## Background

Since the emergence of the severe acute respiratory syndrome coronavirus (SARS-CoV-2) in 2019 there have been intense efforts to understand the natural history of COVID-19 disease and transmission characteristics. The risk of nosocomial transmission has been well reported and personal protective equipment (PPE) requirements for healthcare workers (HCWs) has generated significant discussion internationally ^1^. The World Health Organization (WHO) recently estimated that between 80 000 and 180 000 HCWs have died of COVID-19 up to May 2021^2^.

Aerosol generating procedures (AGPs) such as non-invasive ventilation (NIV) have been considered essential in providing respiratory support for patients with severe COVID-19 and in reducing the need for invasive ventilation which is associated with higher morbidity and mortality, others such as High Flow oxygen have now been shown to offer no benefit in comparison with conventional oxygen^3–5^. Despite improved patient outcomes, AGPs came under increased scrutiny during the severe acute respiratory syndrome (SARS) outbreak of 2003, when observational data suggested AGPs increased the risk of healthcare workers (HCW) acquiring infection^6^. Hesitancy in adopting NIV, high flow oxygen or performing other AGPs may be attributable to perceived risk to HCWs or limited availability of resources thought to be pre-requisite such as negative pressure ventilation and higher-grade PPE^7^. In addition, there is a lack of consensus as to what classes as an AGP and international guidelines (such as WHO, the Centers for Disease Control (CDC) or UK Health Security Agency (UK HSA)) are based on expert opinion due to a lack of solid evidence base.

Air sampling studies in healthcare settings have reported a broad range of frequencies of detection of SARS-CoV-2. Multi-centre environmental sampling performed by UK HSA in 2020 in 8 UK hospitals identified SARS-CoV-2 RNA in 4 of 55 air samples collected <1m from patients^8^. Other studies have reported rates of 0-72%^9,10^ PCR positivity in air samples. More recent environmental sampling within the RECOVERY-RS (Randomised Evaluation of COVID-19 Therapy, Respiratory Support) study included air samples collected during administration of continuous positive airways pressure (CPAP), high flow nasal oxygen (HFNO) and supplemental oxygen. Of 90 air samples from 30 patients 14% were positive for at least one target on PCR and no statistical difference in frequency of positivity was identified between the three groups (CPAP, HFNO and other oxygen)^11^. In healthy volunteers, two studies have demonstrated that coughing, speaking or heavy breathing generate significantly higher volumes of detectable respiratory particles than use of CPAP or HFNO^12,13^.

However, confirmation of the significance of SARS-CoV-2 in the air in hospitalised settings has not been definitive due to the absence of routine air sampling in a prospective COVID-19 patient cohort, with the WHO calling for further studies on the risk of SARS-CoV-2 release during AGPs^7^. A range of approaches have been adopted to investigate the relative risk of aerosol transmission in comparison to other modes like fomite, direct contact or close-range droplet^14^. Further, there is a recognized need to identify which precautions would be most effective in mitigating risks, to better protect communities and healthcare services including vulnerable patients and staff.

This study addresses the question of whether there is an increased likelihood of detecting aerosolized SARS-CoV-2 RNA in the rooms of COVID-19 patients undergoing AGPs than in the rooms of patients requiring oxygen at flow rates of less than 15L/min via mask or nasal cannulae, or not requiring any supplemental oxygen. For the purposes of this study, AGPs are defined as those listed by UKHSA for which specific guidance on infection prevention and control (IPC) measures is provided in the UK (UKHSA 2022) ^15^ (Figure 1).

**Figure 1:** List of procedures defined as aerosol generating procedures (UKHSA) • *Tracheal intubation and extubation • Manual ventilation • Tracheotomy or tracheostomy procedures (insertion or removal) • *Bronchoscopy • Dental procedures (using high speed devices, for example ultrasonic scalers/high speed drills) • *Non-invasive ventilation (NIV); bi-level positive airway pressure ventilation (BiPAP) and continuous positive airway pressure ventilation (CPAP) • *High flow nasal oxygen (HFNO) • High frequency oscillatory ventilation (HFOV) • Induction of sputum using nebulised saline • Respiratory tract suctioning • Upper ear, nose, and throat (ENT) airway procedures that involve respiratory suctioning • *Upper gastro-intestinal endoscopy where open suction of the upper respiratory tract occurs beyond the oro-pharynx • High speed cutting in surgery/post-mortem procedures if respiratory tract/paranasal sinuses involved *indicates procedures included in this study

## Methods

### Setting

Liverpool University Hospitals NHS Foundation Trust have two acute tertiary referral hospital sites within Liverpool, UK (Royal Liverpool University (RLUH) and Aintree University Hospitals (AUH)) that have each managed large cohorts of COVID-19 patients. CPAP, non-invasive ventilation and use of high-flow oxygen were rapidly adopted as therapeutic interventions in Liverpool ^16^ but are listed as AGPs. AGPs were conducted in areas with enhanced ventilation, including pre-existing negative pressure rooms, newly installed temporary systems providing ventilation or areas such as theatre or intensive care. Together with UK-HSA, who had already undertaken air and environmental sampling in hospitals ^8^ we continued to collect air samples from the rooms of patients hospitalised with COVID-19.

Prospective air sampling was performed around inpatients with a recent diagnosis of SARS CoV-2 and in community ambulatory cases (n=13) admitted to the clinical research unit for screening or treatment as part of an early phase trial. Samples included in this interim analysis were collected between November 2020 and October 2021, and the dominant circulating variants in the UK during this period were B.1.1.7 Alpha (PANGO lineage B.1.1.7) and Delta (PANGO lineage B.1.617.2)^17^. Locations sampled include high dependency units, NIV units, critical care units, infectious diseases, respiratory and general wards, and emergency departments. No additional biological samples or clinical information were collected from the patients whose rooms was sampled. No changes to behaviour or clinical care were requested from staff or patients during sampling periods, staff and patients were aware that sampling was taking place. Samples were categorised as either 1) room samples; patients not undergoing any respiratory treatment 2) oxygen; patients undergoing oxygen at flow rates of less than 15L/min via nasal cannulae/mask or 3) AGP; patients undergoing an AGP.

### Sample collection and processing

To collect air samples, we used the Sartorius MD8 AirPort portable air sampler with gelatin membranes (Sartorius, Germany). To ensure aerosols were captured and to minimize the collection of large droplets, the sampler was placed at approximately 1.5 meters^18^ and the sampler faced away from the participant. When sampling at a participant’s bedside, the sampler was typically placed on a hospital chair or table. At each sampling timepoint 1000L of air was collected at a flow rate of 40 L/min.

Once completed the gelatin membrane was removed from the sampler using clean gloves, and placed in a sterile 50ml Falcon tube (Corning Science, United States). Samples were transported to the Biological Safety level 3 (BSL3) laboratories at the Liverpool School of Tropical Medicine within 2 hours of sample collection. The gelatin membranes were immediately suspended in 10ml Dulbecco’s modified Eagle’s medium (DMEM) with 10% fetal bovine solution (FBS; both ThermoFisher, USA) and 50 units per ml of penicillin/streptomycin (Gibco, US). Samples were vortexed and then stored overnight at 4 °C to allow the membrane to fully dissolve without impacting the stability of SARS-CoV-2^19^.

### RNA extraction, PCR and cell culture

Following overnight storage, samples were brought to room temperature by incubation for up to two hours. 140µl (1.4%) of each sample was then taken for RNA extraction. Viral RNA was extracted using the QIAamp viral RNA mini kit (Qiagen, Germany) following the manufacturer’s instructions with an internal RT-qPCR extraction control incorporated before the lysis stage (ThermoFisher, USA). Once extracted samples were taken immediately for downstream application and stored at 4 °C during PCR setup.

For SARS-CoV-2 RT-qPCR detection, 10µl of extracted RNA was tested using the TaqPath™ COVID-19 CE-IVD RT-PCR assay (ThermoFisher, USA) that detects three different SARS-CoV-2 genomic regions (ORF1ab, N gene, S gene), on the QuantStudio 5 platform (ThermoFisher, USA). The PCR was deemed to be positive if one SARS-CoV-2 target and the internal control amplified, this differs from the manufacturer instructions whereby two SARS-CoV-2 targets must amplify however this is for clinical samples rather than for environmental samples. We do not infer viability of the viral RNA we detected and thus the presence/absence of viral RNA is recorded here. In instances of low Ct values the third target can often be out competed and thus viral load calculations have not been appropriate using this assay. Samples were negative if only the internal extraction control amplified.

Cell culture was attempted using a methodology adapted from Edwards et al ^20^. For samples identified as the Alpha variant, based on S gene deletion, a 5-day incubation post inoculation was used due to the variant’s slower growth rate and smaller plaque sizes in Vero E6 cells ^21^. For all other samples a 3-day post inoculation incubation was utilized, as is routine laboratory procedure. After three passage events, samples were fixed with 10% formalin, stained using crystal violet and visually inspected for viral plaques.

### Statistical Analysis

All data handling and manipulation was performed through R 3.6.3^22^. Binomial generalized linear models (GLM) were performed to assess the ability of procedure to detect a positive result, with time since infection, cycle threshold (Ct) value and location of patient included as co-variates. Chi-Square test was performed to estimate the effect of procedure on detecting a positive result in isolation from other factors. Additionally one-way ANOVA tests were performed to assess quantitative differences between Ct values among different methods for each gene.

#### Ethics and Approvals

This study was performed in conjunction with UK HSA as part of the COVID-19 clinical response. In line with UK Health Resource Authority guidance this was performed as a service evaluation and consisted of environmental sampling and simplified and anonymized routinely collected patient information. The environmental sampling process was explained to patients and verbal consent obtained.

## Results (450)

A total of 219 air samples were collected during this study from bed spaces of 199 individuals (with some patient sampled at more than one timepoint) either on room air only (n=67), receiving supplemental oxygen (n=65) or undergoing an AGP (n=87) (Table 1).

**Table 1:** Procedures sampled, and frequency of positive results observed.

| Aerosol generating procedures |  | Non-aerosol generating procedures |  |
| --- | --- | --- | --- |
| Procedure | percent positive (n/N) | Procedure | percent positive (n/N) |
| CPAP hood or mask | 19.2% (10/52) | Room air | 24.6% (17/69) |
| BiPAP | 0% (0/3) | Oxygen - Nasal cannulae | 28.6% (8/28) |
| Hi Flow oxygen (mask or nasal) | 14.3% (3/21) | Oxygen – Venturi mask | 33.3% (9/27) |
| Intubation | 100.0% (1/1) | Oxygen – NRB | 20.0% (1/5) |
| Bronchoscopy | 100.0% (2/2) | Other/Not known | 75% (3/4) |
| Endoscopy | 50.0% (1/2) |  |  |
| All | 20.9% (17/81) |  | 28.6% (38/133) |

CPAP Continuous positive airway pressure. BiPAP Bilevel Positive Airway Pressure. NRB Non-rebreathe.

The median number of days since onset of symptoms was 7 (Table 2). In total, 24.7% (n=55, CI95% 19.4-30.8%) were positive for SAR-CoV-2 viral RNA (Appendix 1). The highest prevalence was identified in the air around patients who were receiving oxygen (32.3%, n=21, CI95% 22.2 to 44.3%) with AGP and room air recording prevalence of 20.7% (n=18, CI95% 14.1 – 33.7%) and 22.3% (n=15, CI95% 13.5 – 30.4%) respectively, as shown in figure 2. The likelihood of a sample being PCR positive was not influenced by procedure (GLM, p >0.05).

**Table 2:** Number of days between onset of symptoms and sample collection within each group.

|  | AGP | Oxygen | Room air | All |
| --- | --- | --- | --- | --- |
| Median number of days since onset of symptoms | 10 (1-27) | 7 (0-42) | 5 (1-12) | 7(0-42) |
| Onset of symptoms unknown | 5 | 2 | 1 | 8 |
| Asymptomatic | 2 | 1 | 17 | 20 |

AGP Aerosol generating procedure.

**Figure 2:**
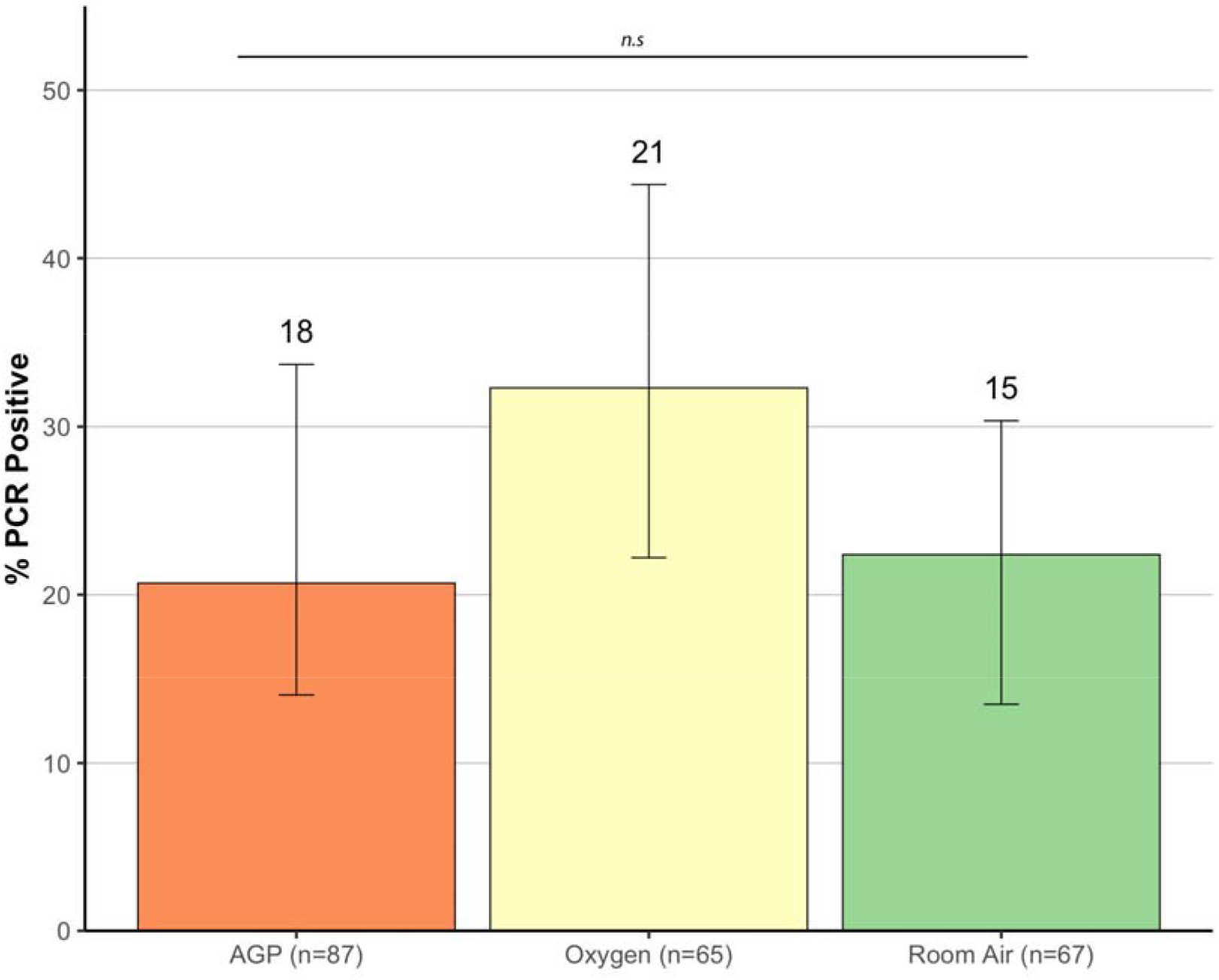
The percentage of samples PCR positive for SARS-CoV-2 viral RNA collected from the rooms of PCR positive patients in two Liverpool Hospitals. There was no statistically significant difference between aerosol generating procedures (AGP), oxygen or room air and the likelihood of detecting viral RNA (GLM, p>0.05). N.s. indicates no statistical difference between the three methods (one-way ANOVA, p>0.05).

We did not detect a difference in Ct values for positive samples between procedure (p>0.05, one-way ANOVA), shown in figure 3. We observed a reduced detection of S gene, indicative of Alpha variant presence, compared to other RT-qPCR genomic targets (ORF1 and N gene). Mean Ct values for AGPs, Oxygen and Room air were 32.18, 33.08 and 33.88 respectively and cell culture for each sample was negative.

**Figure 3:**
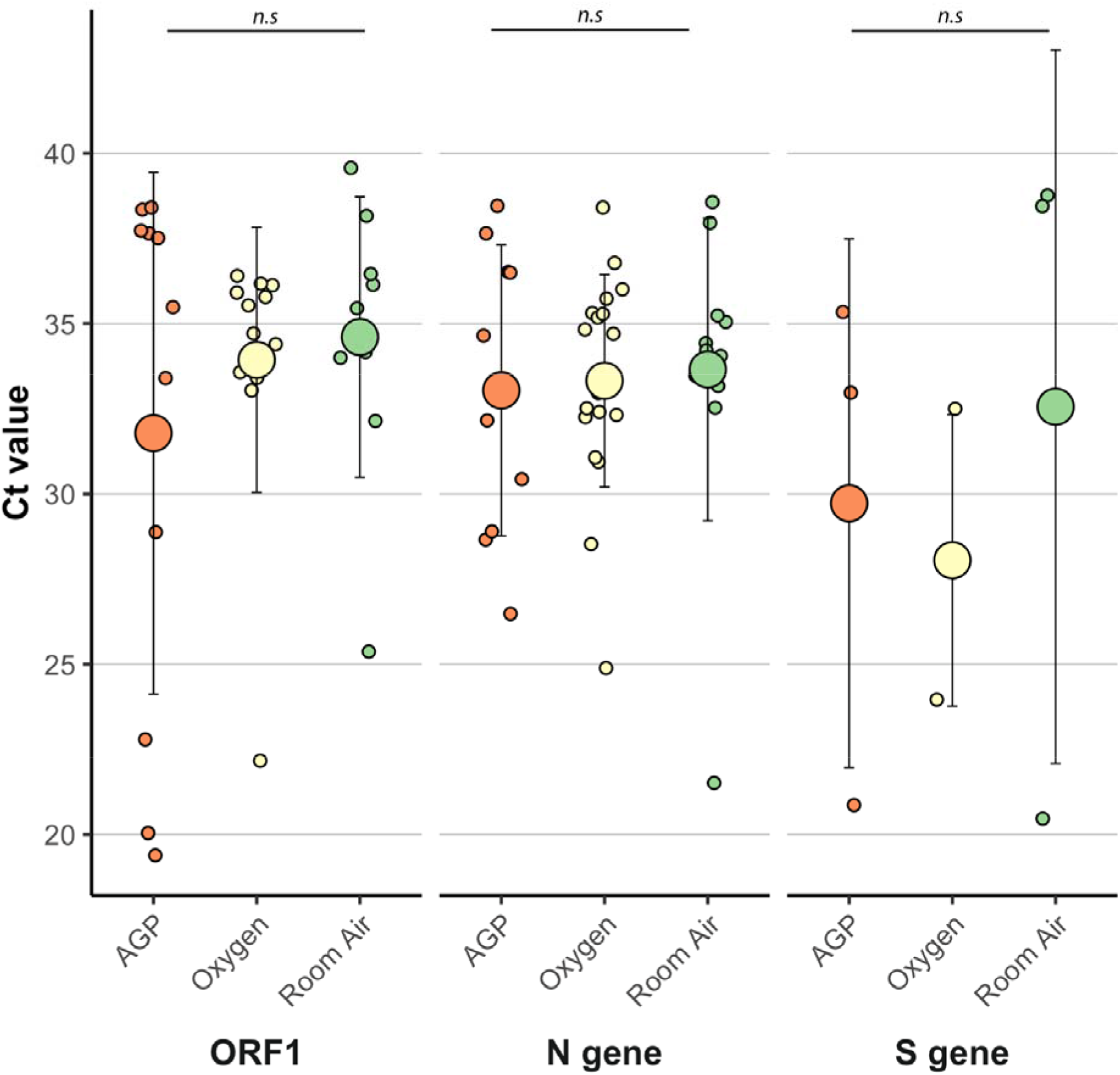
The cycle threshold (Ct) value of SARS-CoV-2 positive samples for each procedure (aerosol generating procedure and oxygen) and room air. The PCR used utilize three distinct regions of the SARS-CoV-2 genome (ORF1, Nucleocapsid (N) gene and spike (S) gene). It is important to note that the alpha variant has significant mutations in the s gene is not detectable, resulting in fewer data points. N.s. indicates no statistical difference between the three methods for each gene (one-way ANOVA, p>0.05).

We did not detect an effect of the number of days since reported onset of symptoms and the frequency of detection of viral RNA or Ct values (GLM, P> 0.05). Further, no association between RT-qPCR results and sampling location e.g., emergency department or critical care unit was identified (GLM, p>0.05). Of note, there was no obvious difference in positive results for the Oxygen or Room air groups when sampling in standard ventilation areas rather than enhanced ventilation areas (RR in Oxygen group 1.22, RR in Room Air group 1.08) (Table 3).

**Table 3:** Type of ventilation, estimated air changes (ACH) and frequency of observed positive PCR results in each group.

| Type of ventilation | Estimated air changes | Intervention |  |  |
| --- | --- | --- | --- | --- |
|  |  | Oxygen | AGP | Room air only |
| Neutral pressure | 6 | 9/25 | 0/0 | 8/39 |
| Negative Pressure | 12 | 10/34 | 15/66 | 4/21 |
| Unknown |  | 2/6 | 0/3 | 0/2 |
| ITU/Theatre | 10 | 0/0 | 3/12 | 0/0 |

ITU Intensive Therapy Unit, AGP Aerosol generating procedure

An additional 16 air samples were taken in areas either occupied or used by patients with confirmed SARS-CoV-2 infection (e.g. bathroom, nursing station), or directly adjacent i.e. considered to be at risk of contamination (appendix 1), SARS-CoV-2 RNA was detected in 5 of these. Culture was negative, and fungal overgrowth was noted in the bathroom samples. Of 4 samples collected in shared patient bathrooms 2 were RNA positive, as were 2 of 8 collected at nursing stations, the remaining positive sample was collected in an ITU bay. Five of the nursing station samples were from stations within ‘red’ zones, 1 from an area adjacent to a red zone, and 2 from ward nursing stations more than 3 metres from COVID-19 patient rooms with separating doors. The Ct values of these 5 samples ranged from 22.4 (S gene) at a nursing station where sample was positive for all 3 genes tested, to 35.19 (for only one gene) for the second positive nursing station sample and patient bathrooms. The positive sample collected in a COVID-19 bay (with the nearest patient intubated) was positive for N gene and ORF1 genes.

## Discussion

The factors that facilitate nosocomial transmission of SARS-CoV-2 remain incompletely understood, including any additional risks posed by AGPs. To improve understanding and inform IPC recommendations we have undertaken the largest air sampling study for SARS-Cov-2 published to date. In this report we provide evidence for the presence of SARS-CoV-2 viral RNA isolated from the air of clinical environments in UK hospitals. In line with previous reports^23,24^, we found Ct values to be high (typically above 30), indicative of low viral loads present. We did not record any significant difference in the observed frequency of viral RNA detection nor viral load between different procedures being undertaken including AGPs. Interestingly, we did observe higher positivity rates in individuals receiving oxygen (flow rates of less than 15L/min via nasal cannulae) than either AGP patients or those without supplementary oxygen although not statistically significant.

The viral dynamics of SARS-CoV-2 in human disease is now well reported with higher viral loads and transmission risk predominantly occurring earlier in disease course. Commonly used AGPs such as CPAP are usually required later in the clinical course with median time from onset of symptoms to development of respiratory failure being 8-12 days^28^. However, we did not see a significant relationship between time since symptom onset and likelihood of viral RNA detection. Infective SARS-CoV-2 virus is rarely identified from clinical samples beyond 9 days from onset of symptoms^26^ and thereby chances of capturing viable virus may be confounded. We were unable to culture samples, likely due to the high Ct values observed, dilutional effect and due to the challenges in culturing virus after potential desiccation during the methods of air sampling. Two reports describe the isolation of infectious SARS-CoV-2 from air sampling. Lednicky et al describe isolation of SARS-CoV-2 virus from hospital rooms used by 2 patients and a vehicle occupied by an infected individual with sequencing of cultured virus matching that of the cases sampled^27,28^. Another challenge in isolating virus from air samples was identified by Lednicky et al as another respiratory virus was found to potentially overgrow SARS-CoV-2 in culture^27^. Santarpia et al described cytopathic effect (CPE), suggested increase in RNA titre on days 3 and 4 of culture and presence of viral proteins on immunofluorescent staining in 1 air sample but could not confirm cultivation of virus^24^. Published aerobiology studies in healthcare settings use a wide array of methods such as filtering air through gelatin, PTFE or PC membranes, using liquid cyclonic sampling such as the Coriolis (Bertin), or impaction e.g., Anderson samplers (Appendix 2). Sampling distances from specified patients sampled varied from 10cm to 3m. Lednicky et al utilised a water condensation collection method (VIVAS) and Santarpia the Sartorius MD8 airport sampler with gelatin membrane filters^24,27^. The diversity of study design and equipment as well as processing and analytical methods presents challenges in comparing the results of different studies. Less than 10 air sampling studies identified through literature search have reported attempted cell culture^10,11,27,29–31^, and the majority have been unable to confirm presence of viable virus despite detection of SARS CoV-2 RNA through PCR^11,29,31^. This is likely attributable to the insensitivity of cell culture, which requires a large inoculum and represents an imperfect model of infection of the human nasopharynx. These challenges are increased when considering environmental samples with low concentrations of detectable viral RNA and the degree of dilution involved in air sampling methods including our own.

It is now accepted that a spectrum of expelled sizes of respiratory particles exist with risk of infection dependent on additional key factors. These include variables relating to the person (e.g., symptoms, duration of illness, individual variation in respiratory particle dispersion) and those related to place (e.g., ventilation, proximity to susceptible individuals) that may be equally or more important than the procedures undertaken. As this was a pragmatic environmental sampling study, detailed patient level data and correlation with clinical SARS CoV-2 samples was not available. This is a recognized limitation as individual characteristics and the viral load detectable on clinical samples may impact likelihood of detection of viral RNA or viable virus in the environment. AGPs were conducted in negative pressure single rooms, pods or bays and bronchoscopy or endoscopy in designated facilities in accordance with Health Technical memorandum 03-01 (including minimum 10 air changes per hour). Those on room air or oxygen were more commonly cared for in rooms with neutral pressure. We also recognise that multiple factors affect the potential impact of infective aerosols including temperature, humidity and ventilation, patient and staff behaviour and pre-existing natural or vaccine mediated immunity.

This study adds to the data of air sampling studies conducted within hospitals caring for COVID-19 patients and offers reassurances to HCW performing AGPs that the presence of SARS-CoV-2 in the air is no different compared with environments where no AGPs are taking place. Our data supports the conclusions drawn from the recent exploratory air sampling study conducted on 30 participants within the RECOVERY RS trial in which CPAP, HFNO and supplementary oxygen via venturi were compared. The observed frequency of detection was not significantly higher in the AGP groups^11^.

Internationally differing lists of AGPs have been produced e.g. including or excluding nebulized therapies or cardiopulmonary resuscitation. In the UK the expert panel responsible for formulating such a list assessed the available evidence as ‘poor quality’^15^. AGPs vary widely and more sampling around a broader range of procedures is required to establish whether there is any evidence of increased risk associated. Any reduction in protections for potentially vulnerable staff or patients regarding AGPs would require more certainty which could only be provided by a more comprehensive evidence base. As we move towards endemicity of COVID-19, it remains imperative to protect HCW and vulnerable patients from nosocomial infections. Appropriate PPE and mask use are essential as SARS-CoV-2 is frequently detectable in the air surrounding positive patients. We believe our data supports the use of FFP2/3 medical masks for HCWs caring for COVID-19 patients, particularly for those with prolonged contact in line with HSE guidelines ^15^. As we move forwards, we must continue to avoid oversimplifying the transmission dynamics of SARS-CoV-2 including into ‘droplet vs aerosol’, as this can lead to fixed IPC recommendations and must now appreciate that the ‘person, place and procedure’ influence transmission risk. Future recommendations must be guided by emerging evidence including ongoing studies to identify transmission differences that may occur with new variants of concern.

## Supporting information

Appendix 1: Details of positive samples Positive samples - procedure, ventilation and Ct values

Details of samples from other areas

Appendix 2: Review of previous air sampling studies Summary of previous air sampling studies in healthcare settings which included details of sampling

Summary of air sampling conducted 1) around specified numbers of SARS-CoV-2 infected patients, 2) around unspecified numbers of infected patients and

## Data Availability

All data produced in the present study are available upon reasonable request to the corresponding author.

## Research in Context

### Evidence before this study

We searched Pubmed (Medline, Web of science (Science citation index-Expanded and Biosis) and Embase for air sampling studies published between January 2020 and November 2021. Selected search terms were “COVID-19”, “SARS-Cov-2”, “air sampler”, “indoor air quality”, “bioaerosol” and “indoor airborne” with 326 results. Studies in which healthcare environments occupied by SARS-CoV-2 infected humans were sampled were eligible. After screening of abstracts and then full texts 37 papers were included.

Frequency of detection of viral RNA in air samples collected from COVID-19 patients varied widely from 0% to 100% and factors affecting likelihood of positivity were not consistently identified. Whilst isolation of infectious virus from air samples is not always attempted, it has only been reported twice but this reflect the challenges of fragile virus capture. Laboratory studies have demonstrated that SARS-CoV-2 may persist for several hours in airborne particles, and epidemiological research has illustrated probable airborne transmission of COVID-19 in settings such as restaurants. Observational data from the SARS-CoV-1 epidemic suggested that aerosol generating procedures (AGPs) presented a greater transmission hazard than other care, that required enhanced respiratory protection. Winslow et al recently reported that in a cohort of 30 patients, that AGPs were not associated with increased likelihood of SARS-CoV-2 viral RNA detection in air compared to supplemental oxygen use. Studies of aerosol generation from procedures such as CPAP and high flow oxygen on healthy volunteers in controlled environments had predicted a low risk of airborne transmission related to such procedures in comparison to cough, speech or singing.

### Added value of this study

As the largest study to date to investigate SARS CoV-2 RNA detection in air samples from rooms of patients undergoing different interventions including AGPS this study contributes significantly to the limited evidence base in this area. We demonstrated that there was no greater frequency of detection of SARS-CoV-2 from the environment of patients undergoing AGPs compared to those on room air or oxygen. This suggests that the risk of aerosol transmission may be equally significant in all areas where COVID-19 patients are cared for and appropriate respiratory precautions should be recommended. This important evidence can provide data to support infection prevention and control guidance in health care settings and inform risk assessments and the use of AGPs in all settings.

### Implications of all available evidence

SARS CoV-2 virus has been identified in the air in a range of healthcare settings where COVID-19 patients are managed. Our findings in combination with those reported by Winslow et al suggest that AGPs do not present a uniformly increased transmission risk. Increased understanding of the natural history of COVID-19 and transmission characteristics of SARS-CoV-2, promote a move from the binary categories of AGP vs non-AGP for respiratory protection, towards a risk assessment based primarily on the Patient and Place, as opposed to just the procedure.

## Appendix 1 Details of positive samples

### Positive samples - procedure, ventilation and Ct values

| Ventilation: Negative pressure (ets. 12 ACH), Neutral or standard ward ventilation (est 6 ACH), ITU and theatre (est. 10 ACH) | Procedure: narrower categories | PCR result | N gene | S gene | ORF1 |
| --- | --- | --- | --- | --- | --- |
| Negative | High flow | Positive | Negative | 20.87 | Negative |
| Negative | CPAP | Positive | 32.16 | Negative | 38.35 |
| Negative | CPAP | Positive | 26.48 | 35.34 | 20.05 |
| Neutral | Room air | Positive | Negative | 38.76 | 25.37 |
| Neutral | Room air | Positive | 37.96 | Negative | 38.16 |
| ITU | CPAP | Positive | Negative | Negative | 37.65 |
| Negative | Nasal cannulae | Positive | 34.83 | Negative | Negative |
| Negative | Room air | Positive | 34.43 | 38.45 | 32.14 |
| Negative | CPAP | Positive | 38.46 | Negative | Negative |
| Neutral | rebreath mask | Positive | 28.53 | Negative | Negative |
| Neutral | Room air | Positive | 33.17 | Negative | Negative |
| Neutral | Nasal cannulae | Positive | 33.03 | Negative | Negative |
| Neutral | Room air | Positive | 34.05 | negative | 34.16 |
| Neutral | Venturi mask | Positive | 32.25 | Negative | 34.71 |
| Neutral | Nasal cannulae | Positive | 36.78 | Negative | 36.13 |
| Neutral | Nasal cannulae | Positive | 32.32 | Negative | Negative |
| Neutral | NK | Positive | 34.7 | Negative | 35.91 |
| Negative | CPAP | Positive | Negative | Negative | 37.51 |
| Negative | CPAP | Positive | Negative | Negative | 22.79 |
| Negative | Venturi mask | Positive | 35.31 | Negative | Negative |
| Neutral | NK | Positive | 32.52 | Negative | 33.04 |
| Negative | Nasal cannulae | Positive | 30.94 | 23.96 | 33.57 |
| Negative | CPAP | Positive | Negative | Negative | 33.4 |
| Theatre | Endoscopy | Positive | Negative | 32.97 | 19.39 |
| Negative | Nasal cannulae | Positive | negative | 27.7 | Negative |
| Negative | Intubation | Positive | 28.66 | Negative | 28.88 |
| Negative | CPAP | Positive | 28.9 | Negative | Negative |
| Negative | Venturi mask | Positive | 24.89 | 32.5 | 22.17 |
| NK | Nasal cannulae | Positive | Negative | Negative | 33.41 |
| Neutral | Room air | Positive | Negative | 20.47 | Negative |
| Neutral | Room air | Positive | 38.57 | Negative | Negative |
| Neutral | Room air | Positive | 33.47 | Negative | 36.14 |
| Negative | CPAP | Positive | 34.65 | Negative | Negative |
| Negative | NK | Positive | 35.73 | Negative | Negative |
| NK | Venturi mask | Positive | 35.18 | Negative | Negative |
| Negative | CPAP | Positive | 37.65 | negative | negative |
| Negative | Venturi mask | Positive | 38.41 | Negative | 36.17 |
| Neutral | Room air | Positive | 35.05 | Negative | 35.45 |
| Negative | Room air | Positive | 35.24 | negative | negative |
| Negative | High flow | Positive | 36.52 | negative | negative |
| Negative | High flow | Positive | 36.5 | negative | negative |
| Neutral | Room air | Positive | 32.53 | negative | negative |
| Neutral | Room air | Positive | Negative | Negative | 36.46 |
| Theatre | Bronchoscopy | Positive | negative | negative | 38.41 |
| Theatre | Bronchoscopy | Positive | negative | negative | 37.73 |
| Negative | Room air | Positive | 34.21 | negative | negative |
| Negative | Room air | Positive | 21.52 | Negative | 39.57 |
| Negative | Room air | Positive | negative | negative | 34 |
| Negative | Venturi mask | Positive | 35.29 | Negative | 35.78 |
| Neutral | Venturi mask | Positive | 31.07 | Negative | 35.54 |
| Negative | CPAP | Positive | 30.43 | Negative | 35.48 |
| Negative | Venturi mask | Positive | 36.01 | Negative | 36.4 |
| Neutral | Nasal cannulae | Positive | 32.41 | Negative | 34.4 |
| Negative | Venturi mask | Positive | 32.96 | Negative | Negative |

NK Not known CPAP Continuous positive airway pressure

### Details of samples from other areas

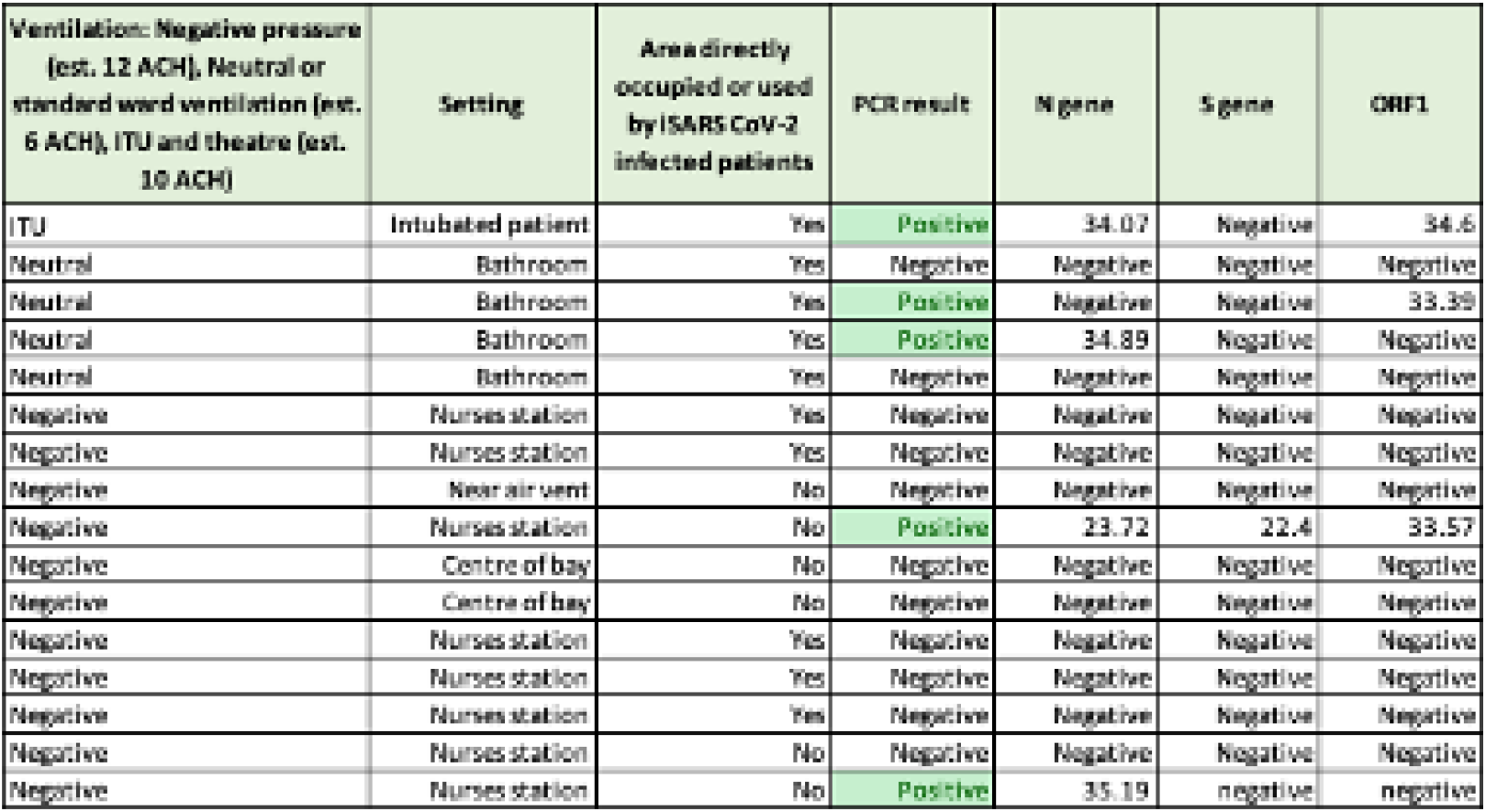

NK Not known CPAP Continuous positive airway pressure Details of samples from other areas
| Ventilation: Negative pressure (est. 12 ACH), Neutral or standard ward ventilation (est. 6 ACH), ITU and theatre (est. 10 ACH) | Setting | Area directly occupied or used by SARS CoV-2 infected patients | PCR result | N gene | S gene | ORF1 |
| --- | --- | --- | --- | --- | --- | --- |
| ITU | Intubated patient | Yes | Positive | 34.07 | Negative | 34.6 |
| Neutral | Bathroom | Yes | Negative | Negative | Negative | Negative |
| Neutral | Bathroom | Yes | Positive | Negative | Negative | 33.39 |
| Neutral | Bathroom | Yes | Positive | 34.89 | Negative | Negative |
| Neutral | Bathroom | Yes | Negative | Negative | Negative | Negative |
| Negative | Nurses station | Yes | Negative | Negative | Negative | Negative |
| Negative | Nurses station | Yes | Negative | Negative | Negative | Negative |
| Negative | Near air vent | No | Negative | Negative | Negative | Negative |
| Negative | Nurses station | No | Positive | 23.72 | 22.4 | 33.57 |
| Negative | Centre of bay | No | Negative | Negative | Negative | Negative |
| Negative | Centre of bay | No | Negative | Negative | Negative | Negative |
| Negative | Nurses station | Yes | Negative | Negative | Negative | Negative |
| Negative | Nurses station | Yes | Negative | Negative | Negative | Negative |
| Negative | Nurses station | Yes | Negative | Negative | Negative | Negative |
| Negative | Nurses station | No | Negative | Negative | Negative | Negative |
| Negative | Nurses station | No | Positive | 35.19 | negative | negative |

## Appendix 2 Review of previous air sampling studies

### Summary of previous air sampling studies in healthcare settings which included details of sampling around aerosol generating procedures (AGPs)

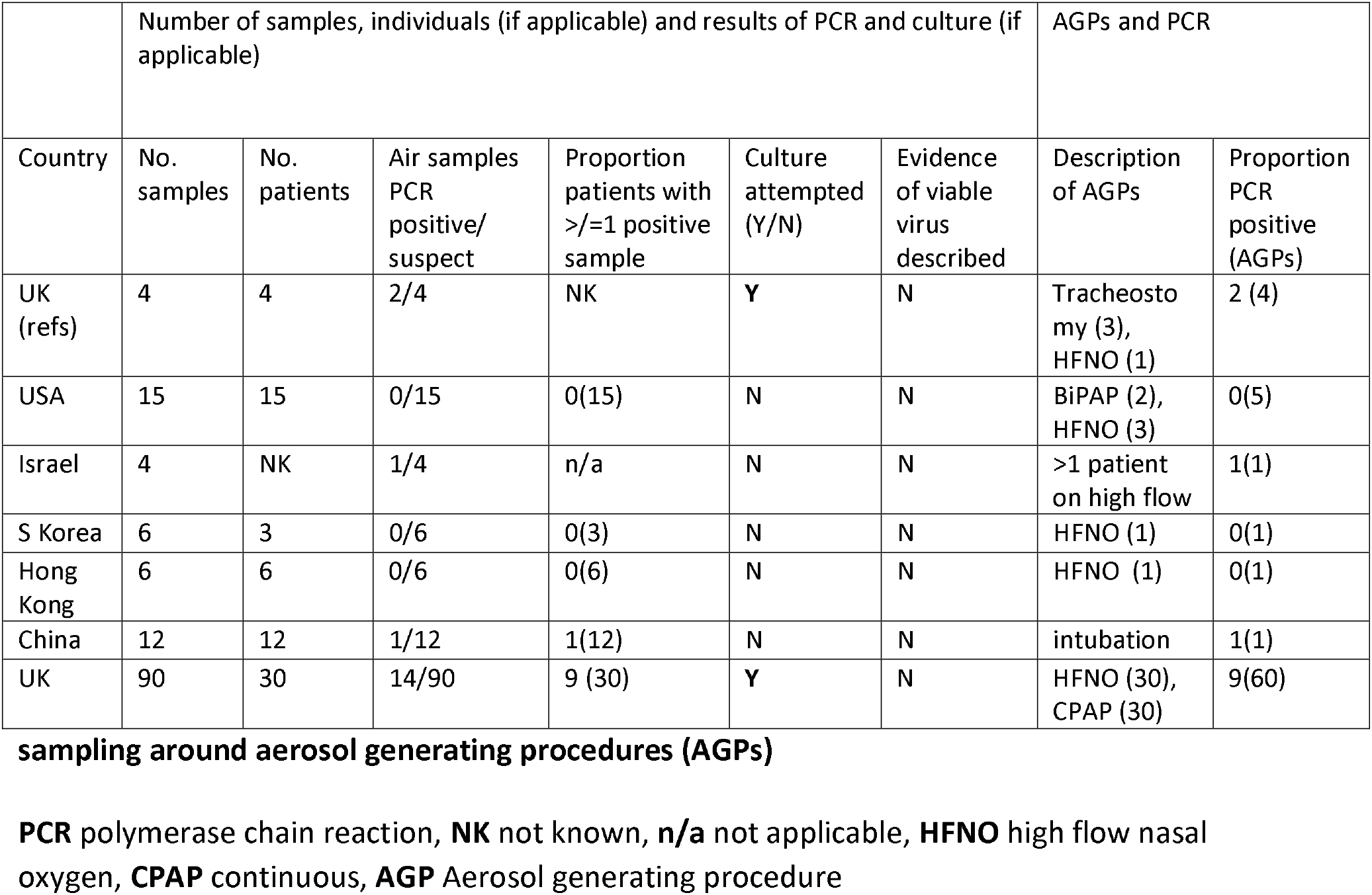

**Summary of air sampling conducted 1**) around specified numbers of SARS-CoV-2 infected patients, 2) around unspecified numbers of infected patients and 3) in areas not occupied by COVID-19 patients from studies where infected patients were also sampled

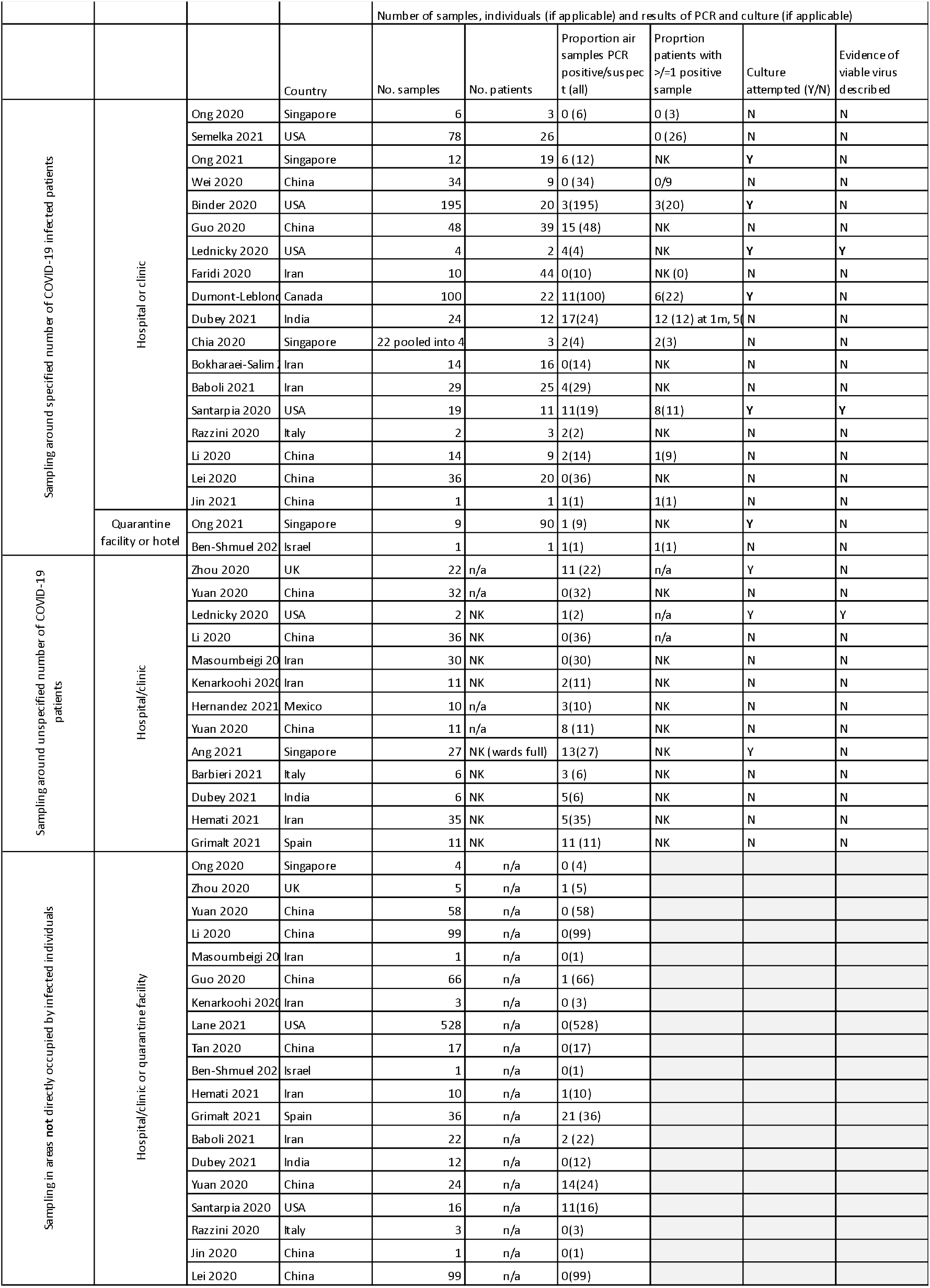

## References

1 Thomas JP, Srinivasan A, Wickramarachchi CS, Dhesi PK, Hung YMA, Kamath A V. Evaluating the national PPE guidance for NHS healthcare workers during the COVID-19 pandemic. Clin Med J R Coll Physicians London 2020; 20: 242–7.

2 World Health Organisation. Health and Care Worker Deaths during COVID-19. 2021. https://www.who.int/news/item/20-10-2021-health-and-care-worker-deaths-during-covid-19 (accessed Oct 21, 2021).

3 Wozniak DR, Rubino A, Tan AL, et al. Positive role of continuous positive airway pressure for intensive care unit patients with severe hypoxaemic respiratory failure due to COVID-19 pneumonia: A single centre experience. J Intensive Care Soc 2022; 23: 27–33.

4 Ashish A, Unsworth A, Martindale J, et al. CPAP management of COVID-19 respiratory failure: A first quantitative analysis from an inpatient service evaluation. BMJ Open Respir Res 2020; 7: e000692.

5 Perkins GD, Ji C, Connolly BA, et al. Effect of Noninvasive Respiratory Strategies on Intubation or Mortality among Patients with Acute Hypoxemic Respiratory Failure and COVID-19: The RECOVERY-RS Randomized Clinical Trial. JAMA - J Am Med Assoc 2022; 327: 546–58.

6 Judson SD, Munster VJ. Nosocomial transmission of emerging viruses via aerosol-generating medical procedures. Viruses. 2019; 11. DOI:10.3390/v11100940.

7 Diaz, Janet; Appiah, John; Askie, Lisa; Baller, April; Banerjee, Anshu; Barkley, Shannon; Bertagnolio, Silvia; Hemmingsen, Bianca; Bonet, Mercedes; Cunningham J. COVID-19 : Clinical management Living guidance. 2021 https://www.who.int/publications/i/item/WHO-2019-nCoV-clinical-2021-1 (accessed Sept 6, 2021).

8 Moore G, Rickard H, Stevenson D, et al. Detection of SARS-CoV-2 within the healthcare environment: a multi-centre study conducted during the first wave of the COVID-19 outbreak in England. J Hosp Infect 2021; 108: 189–96.

9 Li YH, Fan YZ, Jiang L, Wang HB. Aerosol and environmental surface monitoring for SARS-CoV-2 RNA in a designated hospital for severe COVID-19 patients. Epidemiol Infect 2020. DOI:10.1017/S0950268820001570.

10 Ang AXY, Luhung I, Ahidjo BA, et al. Airborne SARS-CoV-2 surveillance in hospital environment using high-flowrate air samplers and its comparison to surface sampling. Indoor Air 2021; : 1–10.

11 Winslow RL, Zhou J, Windle EF, et al. SARS-CoV-2 environmental contamination from hospitalised patients with COVID-19 receiving aerosol-generating procedures. Thorax 2021; : thoraxjnl-2021-218035.

12 Wilson NM, Marks GB, Eckhardt A, et al. The effect of respiratory activity, non-invasive respiratory support and facemasks on aerosol generation and its relevance to COVID-19. Anaesthesia 2021; 76: 1465–74.

13 Hamilton FW, Gregson F, Arnold DT, et al. Aerosol emission from the respiratory tract: an analysis of relative risks from oxygen delivery systems. medRxiv 2021; : 2021.01.29.21250552.

14 Nissen K, Krambrich J, Akaberi D, et al. Long-distance airborne dispersal of SARS-CoV-2 in COVID-19 wards. Sci Rep 2020; 10: 1–9.

15 UKHSA. Infection prevention and control for seasonal respiratory infections in health and care settings (including SARS-CoV-2) for winter 2021 to 2022. Updated 17 January 2022. 2022. https://www.gov.uk/government/publications/wuhan-novel-coronavirus-infection-prevention-and-control/covid-19-guidance-for-maintaining-services-within-health-and-care-settings-infection-prevention-and-control-recommendations#transmission-based-precautions.

16 Nightingale R, Nwosu N, Kutubudin F, et al. Is continuous positive airway pressure (CPAP) a new standard of care for type 1 respiratory failure in COVID-19 patients? A retrospective observational study of a dedicated COVID-19 CPAP service. BMJ Open Respir Res 2020; 7: 8–10.

17 Hadfield J, Megill C, Bell SM, et al. NextStrain: Real-time tracking of pathogen evolution. Bioinformatics 2018; 34: 4121–3.

18 World Health Organization. Modes of transmission of virus causing COVID-19: implications for IPC precaution recommendations. Geneva World Heal Organ 2020; : 19– 21.

19 Chin AWH, Chu JTS, Perera MRA, et al. Stability of SARS-CoV-2 in different environmental conditions. The Lancet Microbe 2020; 1: e10.

20 Edwards T, Kay GA, Aljayyoussi G, et al. SARS-CoV-2 Transmission Risk from sports Equipment (STRIKE). medRxiv 2021; : 2021.02.04.21251127.

21 Jeong GU, Yoon GY, Moon HW, et al. Comparison of plaque size, thermal stability, and replication rate among SARS-CoV-2 variants of concern. Viruses 2022; 14: 4–8.

22 R Core Team. R: A language and environment for ## statistical computing. 2017.

23 Chia PY, Coleman KK, Tan YK, et al. Detection of air and surface contamination by SARS-CoV-2 in hospital rooms of infected patients. Nat Commun 2020; 11. DOI:10.1038/s41467-020-16670-2.

24 Santarpia JL, Rivera DN, Herrera VL, et al. Aerosol and surface contamination of SARS-CoV-2 observed in quarantine and isolation care. Sci Rep 2020; 10. DOI:10.1038/s41598-020-69286-3.

25 Li X, Ma X. Acute respiratory failure in COVID-19: Is it ‘typical’ ARDS? Crit Care 2020; 24: 1–5.

26 Cevik M, Kuppalli K, Kindrachuk J, Peiris M. Virology, transmission, and pathogenesis of SARS-CoV-2. BMJ 2020; 371: 1–6.

27 Lednicky JA, Lauzard M, Fan ZH, et al. Viable SARS-CoV-2 in the air of a hospital room with COVID-19 patients. Int J Infect Dis 2020; 100: 476–82.

28 Lednicky JA, Lauzardo M, Alam MM, et al. Isolation of SARS-CoV-2 from the air in a car driven by a COVID patient with mild illness. Int J Infect Dis 2021; 108: 212–6.

29 Zhou J, Otter JA, Price JR, et al. Investigating Severe Acute Respiratory Syndrome Coronavirus 2 (SARS-CoV-2) Surface and Air Contamination in an Acute Healthcare Setting During the Peak of the Coronavirus Disease 2019 (COVID-19) Pandemic in London. Clin Infect Dis 2021; 73: e1870–7.

30 Binder RA, Alarja NA, Robie ER et al. Environmental and aerosolized severe acute respiratory syndrome coronavirus 2 mong hospitalized coronavirus disease 2019 patients. J Infect Dis 2020; 222: 1798–806.

31 Dumont-Leblond N, Veillette M, Mubareka S, et al. Low incidence of airborne SARS-CoV-2 in acute care hospital rooms with optimized ventilation. Emerg Microbes Infect 2020; 9: 2597–605.

