## Appendix 1: Details of positive samples Positive samples - procedure, ventilation and Ct values for "The aerobiology of SARS-CoV-2 in UK hospitals and the impact of aerosol generating procedures"

**Positive samples - procedure, ventilation and Ct values**

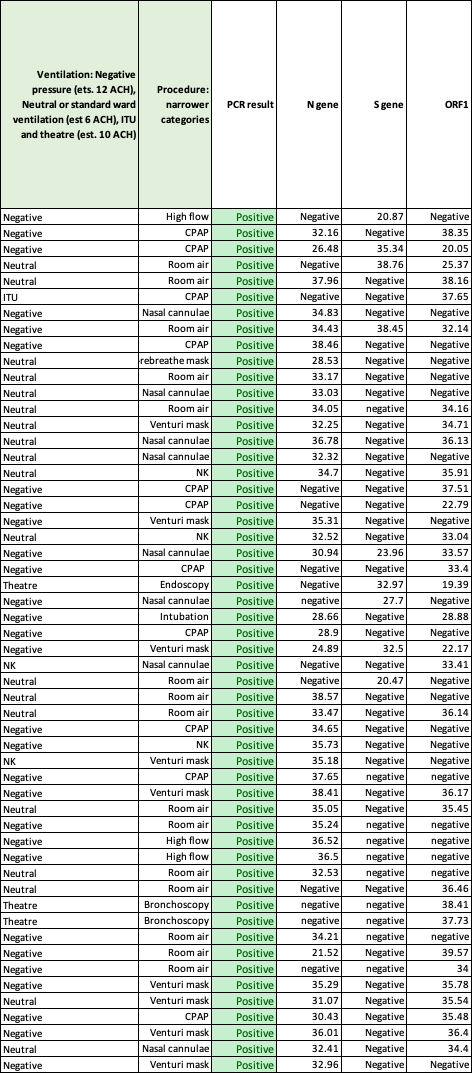

NK Not known CPAP Continuous positive airway pressure
