## Supplementary figures and images for "The aerobiology of SARS-CoV-2 in UK hospitals and the impact of aerosol generating procedures"

### Details of samples from other areas

**Details of samples from other areas**

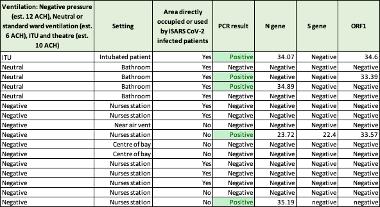
