## Appendix 2: Review of previous air sampling studies Summary of previous air sampling studies in healthcare settings which included details of sampling for "The aerobiology of SARS-CoV-2 in UK hospitals and the impact of aerosol generating procedures"

**Summary of previous air sampling studies in healthcare settings which included details of sampling around aerosol generating procedures (AGPs)**

|  | Number of samples, individuals (if applicable) and results of PCR and culture (if applicable) | | | | | | AGPs and PCR | |
| --- | --- | --- | --- | --- | --- | --- | --- | --- |
| Country | No. samples | No. patients | Air samples PCR positive/ suspect | Proportion patients with >/=1 positive sample | Culture attempted (Y/N) | Evidence of viable virus described | Description of AGPs | Proportion PCR positive (AGPs) |
| UK (refs) | 4 | 4 | 2/4 | NK | **Y** | N | Tracheostomy (3), HFNO (1) | 2 (4) |
| USA | 15 | 15 | 0/15 | 0(15) | N | N | BiPAP (2), HFNO (3) | 0(5) |
| Israel | 4 | NK | 1/4 | n/a | N | N | >1 patient on high flow | 1(1) |
| S Korea | 6 | 3 | 0/6 | 0(3) | N | N | HFNO (1) | 0(1) |
| Hong Kong | 6 | 6 | 0/6 | 0(6) | N | N | HFNO (1) | 0(1) |
| China | 12 | 12 | 1/12 | 1(12) | N | N | intubation | 1(1) |
| UK | 90 | 30 | 14/90 | 9 (30) | **Y** | N | HFNO (30), CPAP (30) | 9(60) |

**PCR** polymerase chain reaction, **NK** not known, **n/a** not applicable, **HFNO** high flow nasal oxygen, **CPAP** continuous, **AGP** Aerosol generating procedure
