## Supplementary material for "The aerobiology of SARS-CoV-2 in UK hospitals and the impact of aerosol generating procedures": Summary of air sampling conducted 1) around specified numbers of SARS-CoV-2 infected patients, 2) around unspecified numbers of infected patients and

**Summary of air sampling conducted** 1) around specified numbers of SARS-CoV-2 infected patients, 2) around unspecified numbers of infected patients and 3) in areas not occupied by COVID-19 patients from studies where infected patients were also sampled

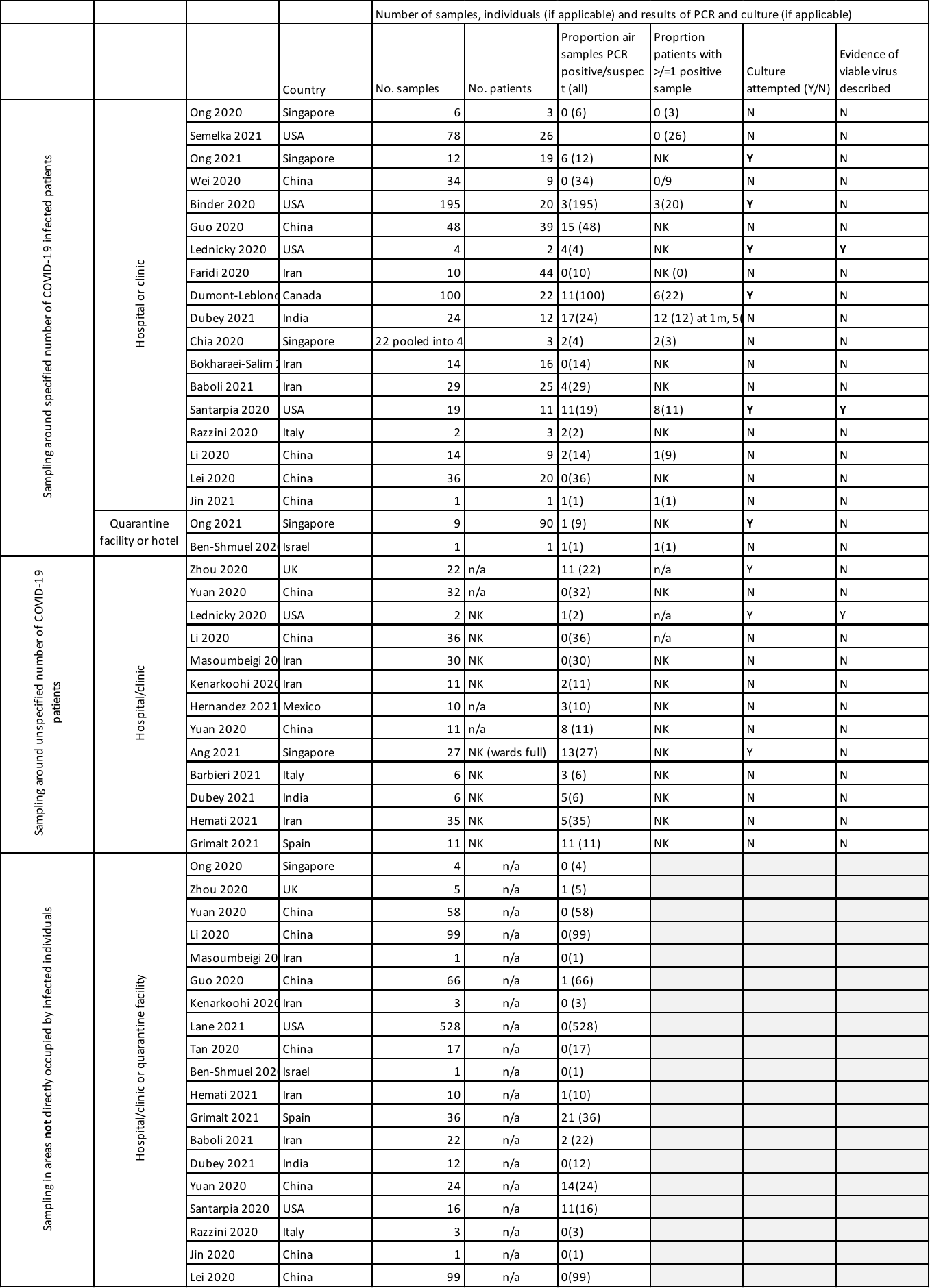
